# Co-development of an acceptance and commitment therapy-based intervention to increase intrinsic motivation of adolescents to change weight: the AIM2Change study

**DOI:** 10.1101/2025.03.04.25323300

**Authors:** Jennifer S Cox, Aidan J Searle, Idoia Iturbe, Gail A Thornton, Ingram Wright, Claire Semple, Ken Clare, Julian P Hamilton-Shield, Elanor C Hinton

**Author notes:** Corresponding author (ECH).

## Abstract

Childhood obesity levels continue to rise, with significant impact on individuals and the NHS. The ‘Complications of Excess Weight’ (CEW) clinics provide support to young people with complications of their weight. Our objective was to co-develop, with young people, a new intervention; AIM2Change, to enable young people to develop their intrinsic motivation to manage weight, using Acceptance and Commitment Therapy (ACT), with a person-centred approach. Young people from the Care of Childhood Obesity (CoCO) clinic in Bristol, UK, were recruited to co-develop this intervention. The study was registered on ISRCTN (ISRCTN16607863). The seven-session, ACT-based intervention was delivered one-to-one, securely online. Qualitative interviews were conducted after each intervention session was delivered. Qualitative data were coded and reviewed regularly to make iterative changes to individual sessions and the overall programme. Fourteen co-developers were recruited, of whom nine completed the co-development process (female=4; median age (IQR)=15(1.5); 4 with a parent; Indices of Multiple Deprivation (IMD) median = 3.5, range=1-10). Iterative changes made during co-development included introducing an earlier focus on eating behaviour and body image, with more practical activities to increase engagement. Thematic analysis of co-developer feedback identified four themes: theoretical understanding; delivery and receipt of therapy; view of strategies and engagement; real world benefits of co-development process. Framework analysis was conducted to map data pertaining to these themes into matrices according to each participant and session. Insights from the co-development process have shaped AIM2Change to optimise the intervention’s value, relevance and acceptability. Findings suggest that AIM2Change meets an unmet need in delivery of current childhood weight management services.

## Introduction

The prevalence of childhood obesity remains alarmingly high(1), with over 340 million children and adolescents classified as overweight or obese worldwide(2). Moreover, among high-income Western countries, English-speaking and Mediterranean regions have the highest levels of obesity in children and adolescents aged 5-19 years(3). Childhood overweight or obesity is associated with many complications, including metabolic (e.g. dyslipidaemia, hypertension, and steatotic liver disease) and psychological (e.g. anxiety and reduced quality of life) comorbidities(4, 5), which are maintained into, or emerge in, adulthood(6). The early onset of obesity that persists into adulthood is associated with greater morbidity, including increased risk of hepatic steatosis and systolic hypertension (7), than obesity that arises in adulthood alone(8). To this is added the financial cost to the NHS(9).

The National Institute for Health and Care Excellence (NICE) guidance recommends multicomponent lifestyle interventions as the preferred option for tackling obesity in childhood and adolescence(10). These interventions promote changes in calorie, nutritional intake and physical activity, which have demonstrable benefits for reducing abdominal obesity(11). There have been numerous governmental schemes and policy changes directed towards reducing obesity in young people (12), yet levels of obesity continue to rise (13). Surgical interventions remain a rarely utilised option(10). However, if healthier lifestyle recommendations have proven unsuccessful, and patients are over 12 years age and/or have comorbidities, the option of using pharmacological treatments is considered. The US Food and Drug Administration (FDA) and the European Medicines Agency (EMA) have approved treatment with subcutaneous Glucagon-Like-Peptide 1 (GLP-1) agonists as an adjunct to lifestyle interventions in adolescents, under certain conditions(14). However, the long-term effects of such medication are unknown and often treatment is restricted to a two-year therapeutic period.

Previous work by our group (15) identified that the young people and their parents attending the CoCO clinic had low levels of self-determination and intrinsic motivation, and were seeking medical-led interventions. Self-determination is known to be important when sustaining lifestyle behaviour change to manage weight (16,17). In a further publication, Cox et al (18) suggested applying Acceptance and Commitment therapy (ACT) to increase self-determination for weight management in young people, because of the way that ACT guides the individual to clarify what intrinsically motivates their behaviour and make commitments to enact these behaviours(19).

In adult weight management, ACT has the strongest evidence amongst the third-generation treatments(20–22) and a recent scoping review of ACT in adolescent weight management showed treatment acceptability, attendance, retention and appraisal were found to be high, whilst weight and quality of life-related outcomes were promising, highlighting the need for further studies in this area(23). A variation of ACT, the DNA-V (Discoverer, Noticer, Advisor-Values) approach(19), offers an evidence-based framework that has been specifically developed for use with young people. It uses ACT processes to teach young people skills to navigate themselves and their social world (24), supporting them to make decisions and changes in their life in ways that can be directly applied to the lifestyle changes required for weight management.

The proceeding work is the next step in realising a novel intervention for intrinsic motivation for weight management services for young people. The objective of this research was to co-develop a DNA-V based intervention, AIM2Change, with young people living with obesity, using the person-based approach (25). This research facilitated co-developers to experience the AIM2Change sessions, and provide real-time evaluations to help to shape the resulting final therapy. PPI contributions from a geographically separate, wider group of young people living with obesity guided the design and implementation of the research study itself, and also shared reflections on the intervention development. The overall aim of this research project was to have a finalised, co-developed intervention that could then be take forward to feasibility testing in a future research trial.

## Materials and methods

### Study design and setting

This study was approved by the UK NHS North-West Research Ethics Committee (ref: 22/NW/0337) and was registered in February 2023 in International Standard Randomised Controlled Trial Number (ISRCTN) database (https://www.isrctn.com/ISRCTN16607863). The study was conducted in the context of a tier-three, paediatric weight management service in Bristol (UK). Recruitment began on 7th December 2022 and ended on 7^th^ June 2023. Additional information regarding the ethical, cultural, and scientific considerations specific to inclusivity in global research is included in the Supporting Information (S1 File).

### Participants

Young people attending the CoCO clinic at Bristol Royal Hospital for Children were approached to collaborate in the intervention development if they (i) were between 11 and 18 years of age and (ii) had a body mass index (BMI) of > 95th percentile. For recruitment, members of the MDT at the CoCO clinic at Bristol Royal Hospital for Children offered the study information pack to young people who needed additional support for weight management beyond the current multidisciplinary treatment in their regular appointments. Next, the ACT therapist with the role of delivering the co-development therapy sessions (JC), contacted the young people and their families to answer any questions they had about the study and their participation. If the young person was interested, written informed consent or assent was obtained online from young people and/or their parents (dependent on age), and appointments were scheduled.

### Procedures

#### PPI

The importance of PPI in the development of novel interventions has been highlighted by the NIHR (26, 27). Accordingly, collaborators with lived experience of obesity, recruited through CEW clinic contacts, the SHINE weight management programme, and social media advertisements, provided input throughout the research process: (i) pre-ethics committee approval (two sessions); (ii) during the data-gathering and analysis stages (two sessions); (iii) design of next study (one session); (iv) discussion of findings and to get feedback on our PPI approach. All PPI sessions were facilitated by our Public Contributor research team member (GT), who has advised and participated throughout the research process, and co-authored this paper. Importantly, there was no overlap between the PPI advisory group members and the young people in the co-development sessions (referred to as participants or co-developers for greater clarity).

The PPI Advisory Group meetings were evaluated using the CUBE methodology(27, 28) to understand perceptions of attendees’ experience in four areas: how they could contribute (one-way to many ways), voice (strong to weak), agenda-setting (public members to organisation) and potential to change the intervention (organisation resistant to intervention changes up to willingness to change). After the meetings, attendees answered each question using a slider under each question from a score of 0 at one end of each dimension up to 1 at the other end. Contributors could also add a comment regarding each dimension. All data were recorded anonymously in a spreadsheet associated with the CUBE link. The mapping of responses on the CUBE could be viewed by all contributors (after logging into the online system).

#### The AIM2Change Intervention

The template intervention was based on the DNA-V model of ACT (24) aiming to increase intrinsic motivation in young people for making and maintaining lifestyle changes by connecting them with their personal values(18). It comprised seven 45-minute sessions (Fig 1) and was delivered on an individual basis (by JC) using the Microsoft Teams online platform: minimising clinic visits, saving time and travel costs. Where required, arrangements were made to provide computer tablets and data packages, and participants were offered a suitable, private, space in which to hold the sessions. Concepts from the model were introduced in each session, along with relevant metaphors and experiential exercises to help the young person understand how to apply the concepts and activities in their lives. The young people were encouraged to practice the skills and techniques in between the sessions in their particular contexts.

**Fig 1.**
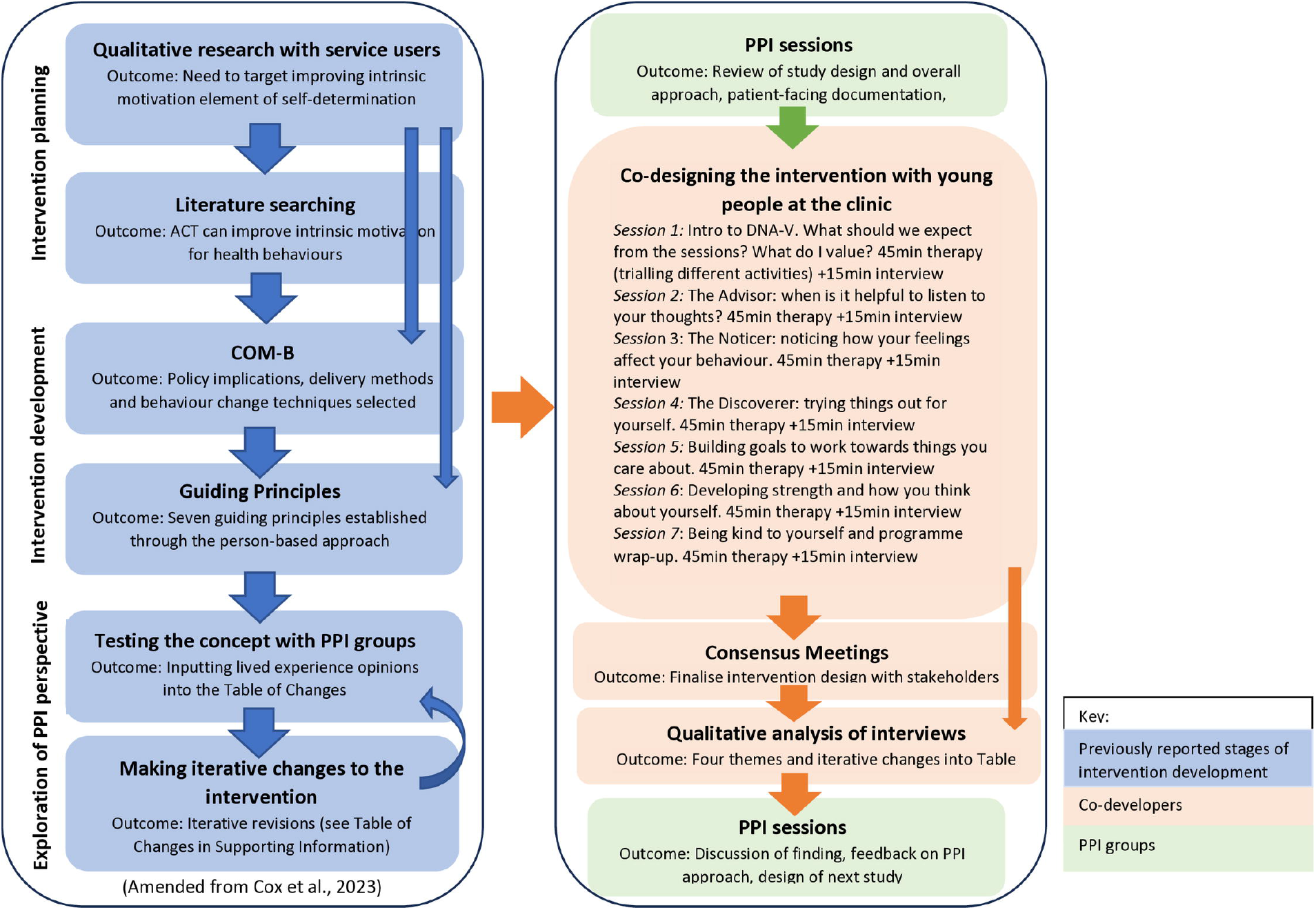
Stages and theoretical framework of AIM2Change intervention development.

**Fig 2.**
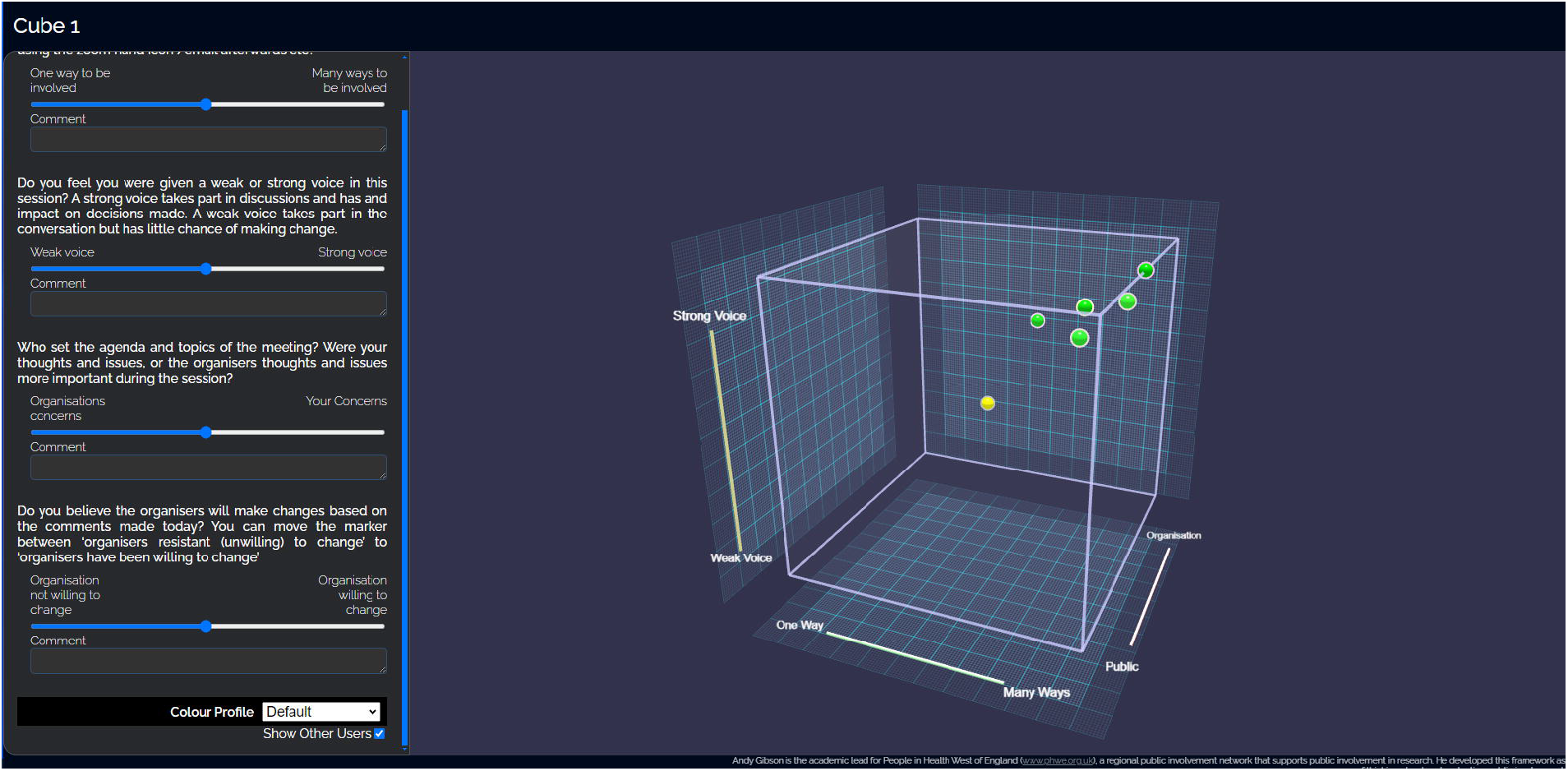
Screenshot of the CUBE. Questions for each dimension appear on the left of the screen with the slider to move the data point in blue. The CUBE appears on the right, with each dimension on a different axis: Contribute on the x□axis, Voice on the y□axis, agenda on the z□axis and change as colour of the data point (yellow is neutral; red is resists change, green is willing to change). The cube can be rotated using the mouse so that contributors can explore the data points in relation to the different dimensions (27, 28).

#### Co-development with Qualitative interviews

The initial therapy framework was iteratively co-developed with the participants, through 15-minute semi-structured interviews. The interviews were conducted by a qualitative researcher (AS) immediately following each therapy session to elicit feedback. The interviews were appraised on an individual basis rather than on aggregated data from multiple participants/sessions, to enable real-time changes to be made. Pertinent feedback, shared during the therapy sessions and the interviews, was compiled into a ‘Table of Changes’ (S1 Table), along with the insights from the PPI Advisory Groups and consensus meetings (see below). Once a point was raised, evidence in support of this suggestion, and opposing thoughts, were collated in the table. The research team (EH, AS, JC) met regularly to discuss the content of the Table of Changes and make iterative changes to the intervention accordingly. The MoScoW (Must do, Should do, Could do, Would like to do) coding scheme(25) was used to classify suggestions. Appropriate changes were also made to the interview topic guide to gather further opinions on the suggestions raised.

#### Consensus meetings

Once all participants had completed the intervention, two consensus meetings were held to discuss the proposed changes and make additional refinements, with the aim of coming to a consensus about the final intervention manual. The first meeting involved participants in the co-development process (young people and their parents if involved), and the second meeting involved members of the clinical MDT, collaborators involved in the research and the management group. Agreed amendments were recorded in the ‘Table of Changes’ (S1 Table).

### Data analysis

#### Coding of Qualitative Interviews and consensus meeting

Inductive coding was used to explore participants’ overall experience of receiving and engagement with the therapy. All interviews were transcribed by AS and II and were managed in NVivo software. Independent ‘open’ coding was undertaken, by AS and II, on a subsample of the interview transcripts (7 of 63), representing four participants: one interview from each of the seven therapy sessions. The initial coding and any discrepancies in coding between the coders were discussed until consensus was reached. A final coding frame was developed jointly, and then applied to the entire dataset by AS. The codes were discussed further by AS, II and EH to inform the development of four themes. The consensus meeting with young people and/or parents was open coded using existing codes developed in the analysis of the ‘Think-aloud’ interviews. (NB: the second consensus meeting was not subjected to coding and analysis, as the focus of that meeting was on the practical delivery of the intervention within the NHS. Insights were captured through inclusion in the Table of Changes, S1 Table).

#### Framework analysis

A framework approach enabled summarising the data pertaining to the four themes in matrices that display the responses from (i) each participant and (ii) each therapy/intervention session attended(29). These coded data from the consensus meeting were also integrated by AS and II into the framework matrices, wherever they were considered in relation to the existing themes (e.g. provided additional insight or perspective to responses derived from the Think-aloud interviews).

## Results

### Patient and Public Involvement in the study

Seventeen young people with relevant personal experience contributed to the study over the course of the overall project, and nine adults, with either personal or parental experience of living with obesity. They came from the South-West of England, Bristol, Sheffield and Birmingham.

### Outcomes from the Patient and Public Involvement process

The advisory group members collectively provided feedback and suggestions on the design and desirability of the research; practicalities of recruitment (including helping with wording of recruitment materials and participant information both for young people and their parents); and offered helpful guidance on how to approach recruitment sensitively. Throughout, the contribution of the advisory group actively influenced the conduct and delivery of the intervention.

The results of the evaluation of Advisory Group members’ experience of their involvement are given in Table 1, which presents the scores allotted to each CUBE dimension(27, 28). In summary, the advisory group evaluated the meetings as interesting, allowing different ways to participate and felt the team were highly likely to make changes resulting from their input.

**Table 1.**
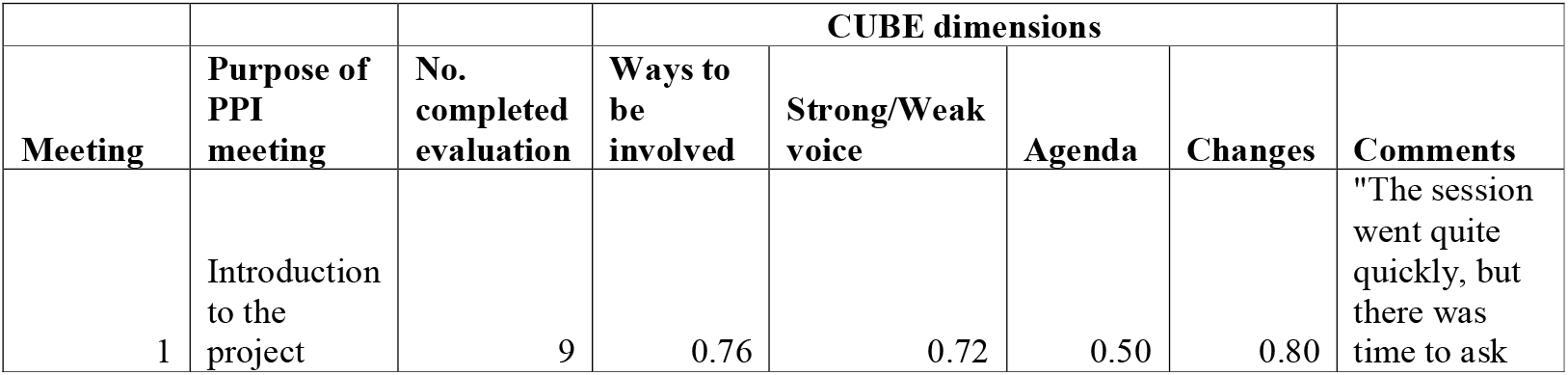
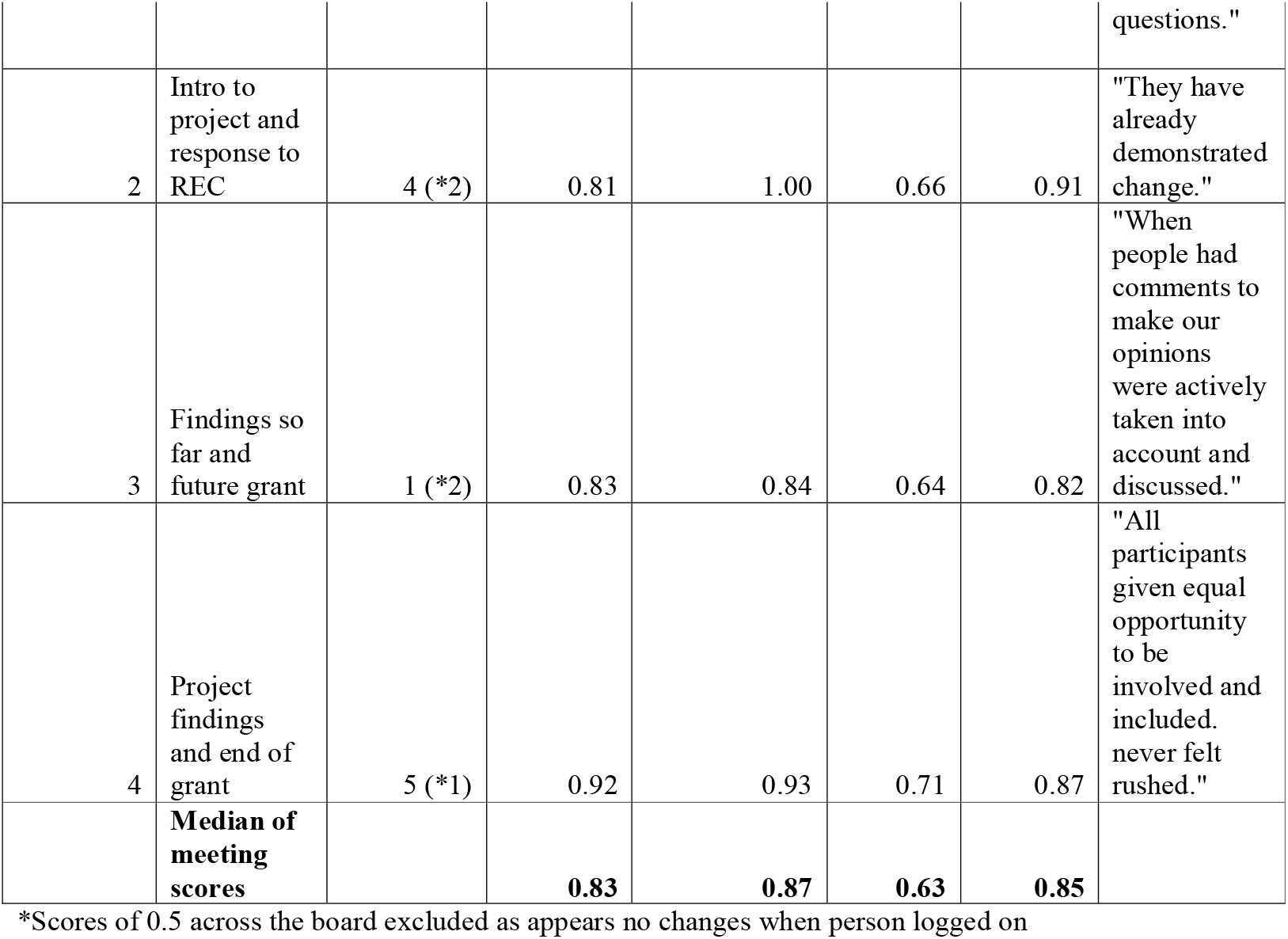
CUBE evaluation of PPI sessions by advisory group members.

|  |  |  | CUBE dimensions |  |  |  |  |
| --- | --- | --- | --- | --- | --- | --- | --- |
| Meeting | Purpose of PPI meeting | No. completed evaluation | Ways to be involved | Strong/Weak voice | Agenda | Changes | Comments |
| 1 | Introduction to the project | 9 | 0.76 | 0.72 | 0.50 | 0.80 | "The session went quite quickly, but there was time to ask questions." |
| 2 | Intro to project and response to REC | 4 (*2) | 0.81 | 1.00 | 0.66 | 0.91 | "They have already demonstrated change." |
| 3 | Findings so far and future grant | 1 (*2) | 0.83 | 0.84 | 0.64 | 0.82 | "When people had comments to make our opinions were actively taken into account and discussed." |
| 4 | Project findings and end of grant | 5 (*1) | 0.92 | 0.93 | 0.71 | 0.87 | "All participants given equal opportunity to be involved and included. never felt rushed." |
|  | **Median of meeting scores** |  | **0.83** | **0.87** | **0.63** | **0.85** |  |

\*Scores of 0.5 across the board excluded as appears no changes when person logged on

### Co-development

#### Participants’ characteristics

Fourteen co-developers were recruited (Fig 3, Table 2), although one did not start due to access difficulties. Of the 13 remaining individuals (six female; mean age, 14.9 years; mean IMD score 4.9), neurodiversity was experienced by two participants with Autism Spectrum Disorder (ASD) or ASD and Attention deficit hyperactivity disorder (ADHD) and seven were receiving a GLP-1 agonist as adjuvant treatment. Eight participants recruited had previous experience of psychological therapy. When asked the question “How would you describe your ethnicity?”, participants used the following terms to describe themselves: Afro-Caribbean (n=1), Black African (n=1), Black British (n=1), Eastern European White (n=1), Pakistani (n=2), Roma Gypsy (n=1), White Romanian (n=1), White (n=2), and White British (n=4).

**Table 2.** Sample characteristics.

|  | Enrolled ppts<br>(*n* = 14) | Completed ppts<br>(*n* = 9) |
| --- | --- | --- |
| Variable | *f*/M (SD) | *f*/M (SD) |
| Female | 6 | 4 |
| Age | 14.9 (1.58) | 14.6 (1.54) |
| IMD | 4.9 (3.16) | 5.2 (3.31) |
| Neurodiversity | 2 | 1 |
| Prescribed GLP-1 Agonist | 7 | 6 |
| Psychological therapy experience | 8 | 5 |

IMD = Indices of multiple deprivation; M = Mean; Ppts = participants; SD = Standard Deviation.

**Fig 3.**
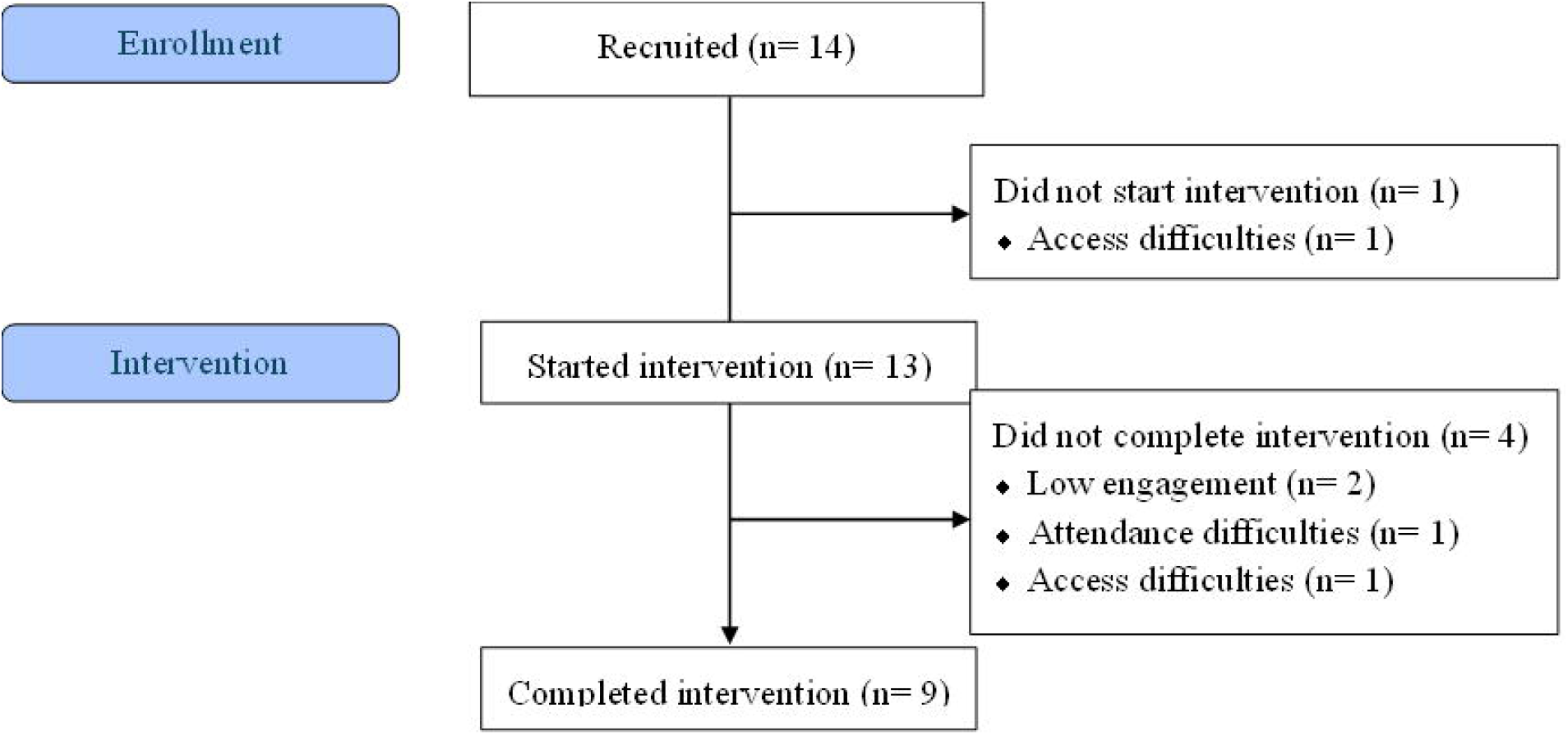
Participant flow diagram. This diagram is prepared in the style of a CONSORT flow diagram (https://www.consort-spirit.org/)

#### Co-development participation

Co-developers who started the intervention attended a mean of 5.1 sessions, and rescheduled sessions twice on average. Of the nine completing all 7 therapy sessions, seven completed all seven interviews, with two participants missing 1-3 interviews. Of the remaining five consenting participants, one completed two interviews, two completed one interview, and two did not complete an interview. In total, 63 interviews were conducted and were analysed as described below. The completed intervention had a total mean duration of 47 days. Parents involved in sessions attended an average of 4.8 sessions.

Two consensus meetings were held online in June and July 2023. All the young people who participated, and their parents, were invited to attend. Six young people and four parents attended, alongside members of the research team (JC, GT, EH). The second meeting was attended by 10 clinical MDT members, collaborators and management group and research team members (including a young PPI member of the management group) to share views, experience and reach consensus on proposed changes.

#### Outcomes from the co-development process

The four themes from the interviews (Table 3) are discussed below together with the key findings documented in the Table of Changes (S1 Table) and the resulting outcomes to implement in the final intervention.

**Table 3.** Thematic structure and coding description.

| Themes | Codes informing theme | Description |
| --- | --- | --- |
| **Theoretical understanding** | Identification and relevance of DNA-V; understanding and relevance of metaphors; assimilation of concepts | Participants' ability to understand how to apply the DNA-V approach to their lives and research team identifying the best examples and metaphors to facilitate this understanding |
| **Delivery and receipt of therapy** | Alignment with therapy aims; openness and receptivity to therapy; focus of sessions; structure and format of sessions; parental involvement; written materials and workbook; continuity and coherence of sessions; uniqueness of the contribution of therapy | Concurrence with aims of the study i.e. co-development. Issues with the structure and format of individual sessions, openness to therapist and suggested approaches / strategies. Practicalities – online or in-person |
| **View of strategies and engagement** | Putting strategies into practice; usefulness of strategies and metaphor; proposed changes to future sessions; satisfaction with session; continuity and coherence of sessions | Extent to which strategies and metaphors are received by participants, and they feel able to, or have already been, put into practice. Evaluating understanding of strategies in context of perceived usefulness and how they relate to their own behaviour |
| **Real world benefits of co-development process** | Reported benefits of attending therapy sessions; view of therapy coming to an end | Self-reported positive outcomes associated with psychological strength and intrinsic motivation, both behavioural (e.g. eating behaviour, setting and appraising goals) relational / social (i.e. communication) outcomes |

#### Qualitative theme: Theoretical Understanding

Through this theme, it emerged that there was no “one size fits all” approach to understanding DNA-V; different activities worked for different people.

> Interviewer: *What was offered for your stress, what strategies were on offer?* Ppt: *Oh, we did try breathing, counted breathing. It was very helpful*. (Ppt 8, Session 5, Male)
>
> *In previous ones* [sessions], *we talked about the Advisor. But it wasn’t about one big thing really. But today we talked about lots of different things. Yeah, we did where you shut your eyes and the leaves on the stream* [therapeutic exercise] *and everything. Yeah, that was quite good*. (Ppt 4, Session 5, Female)

Whilst the videos and generic explanations of the DNA-V concepts were well received, young people valued the support of the therapist guiding them to understand and apply the DNA-V concepts to their own lives, values and experiences:

> *After that* [video], *we started talking about those three parts of the brain and that went on for the rest of the time, talking about examples of it. OK. It was really well explained. At first, I was thinking, well, how do these relate to how people think? Because I didn’t think how well it could be simplified into three different aspects. But eventually realised it does fit perfectly, whether you’re, like, a little voice in your head, or what. You’re just noticing things. It was familiar to me, I never had quite a name for it, but it was recognisable*. (Ppt 7, Session 1, Male)
>
> *We went back on to what we did last week, which was starting off with the DNA. And then we added the “V” in the middle, so it was like a geometric shape type thing. And it was quite helpful because we looked at one and then we were like, OK, Ohh, how can you apply it?* (Ppt 1, Session 2, Female)

The space for self-reflection also supported young people to link the programme with their own experiences:

> *There’s, like, a good amount of self-reflection, I suppose, and sort of placing emotions on certain memories. It makes you feel stuff like that*. (Ppt 2, Session, Female)

##### Outcome

The manual will contain ‘core content’ that carries the fundamental DNA-V content followed by several different, flexible, exercises that explore the concepts in slightly different ways. Together the therapist and patient will experiment to see which activities help them to understand and embed the content in the context of their life and experience. Therefore, all patients will be guided using the same therapeutic principles led by DNA-V but will not experience an identical programme.

#### Qualitative theme: Delivery and Receipt of therapy

Throughout early PPI work, the whole-person approach to therapy was commended, as opposed to an approach that solely looked at issues around weight. However, during the intervention co-development itself, some young people requested that discussions of weight, eating and self-image were introduced earlier in the therapy to have time to expand on the issues raised. However, other young people noted that the topic was difficult to discuss, and that balance was important:

> *It’s a hard thing for me to talk about. I just don’t feel comfortable with it, but it’s sort of like putting in a bit of it to every single aspect that we talk about, that sort of the right amount rather than being constantly talking about it* (Ppt 7, Session 5, Male)

The therapeutic relationship was deemed important for therapeutic success, and building rapport was often successful:

> *I think the environment when I was talking to JC* [therapist] *was quite comfortable and welcoming. I think at the start of the session I was quite stressed out and now I’m feeling quite calm through the techniques that JC taught me and that we went through today. Well, I guess I never really talked to someone about how I felt*. (Ppt 1, Session 1, Female)
>
> *First time ever having therapy, so I was a bit nervous about what to expect. I was quite shocked It was very well explained. I think it was structured and planned out quite well*. (Ppt 7, Session 1, Male)

Not all began the sessions feeling able to engage:

> *I don’t like doing it* [talking] *with people like strangers. I just don’t feel comfortable. But even if I do know the person, it’s not like it’s guaranteed that I’ll tell them everything, like. Weird in a way to say everything you feel whenever you feel and whatnot*. (Ppt 12, Session 1, Male)

However, resistance to engagement began to dissipate over the course of the sessions:

> *I mean, it’s still, it’s not, like, the most enjoyable thing ever. So still quite boring, but it’s not as bad as I thought*. (Ppt 12, Session 3, Male).

The choice, in the design, of online therapy was made initially for pragmatic reasons, to reduce the time commitment and cost to the co-developers of traveling to the hospital weekly. Feedback suggested that this did not affect the rapport:

> [I’ve] *had therapy before with someone online, and it never really flowed*… *Whereas this flows, it absolutely flows. It feels like you’re in-person*. (Ppt 4, Session 3, Female)
>
> Ppt 7: *I like doing it on Zoom as well, I felt quite safe and secure*. Mother: *He was in his room. He was on his territory* (Ppt 7, Consensus meeting, Male)

However, several young people could not find a private area at home to receive the therapy, and experienced frequent interruptions:

> Interviewer: *What about your situation at home, is it private enough for you?*
>
> Ppt: *Yeah. Go[a]way* [shouting to Brother]. (Ppt 9, Session 7, Male)

This sibling interaction did not seem to affect this young person’s experience of the therapy but is an important consideration for the therapist when session planning.

The co-developers felt that the approach they received, to have one-to-one sessions was optimal:

> *I don’t really like the idea of group therapy because everyone has different opinions on things and they have different things that they want to talk about, and, depending on how long the group therapy lasts for, I feel like not everyone gets to have a say*. (Ppt 1, Session 7, Female).

The earlier learnings regarding tailoring the explanations to the individual’s circumstances, and adapting activities, also lend themselves to one-to-one therapy.

The intervention design intentionally allowed participants to choose the level of involvement their parents had in the therapy, and young people reflected on this favourably:

> *I wouldn’t be as open as I probably am. I think if I was to have my parents* [it] *would probably be a big argument*. (Ppt 1, Session 7, Female) *I like it better just me* (Ppt 10, Session 7, Male)
>
> *I personally like my mum being with me, but because I’m quite open with those sort things. But I guess it depends on what sort of relationship you have with your parents. If you’re quite open with them or not, in a way. So, for some people it can be good, but for some people they may not like it*. (Ppt 4, Consensus meeting, Female)

Finally in this theme, the young people concluded that they would like to have had more therapy sessions than the seven they received:

> *I got the impression that it took a few sessions to kind of get going and then when it was really getting good it was finished*. (Ppt 2, Female) *So yeah, I think a bit longer might have, you know, that it might have been quite good in the next four sessions*. (Ppt 2, Female)
>
> *I think that it did take a couple of sessions to get used to what we were, like, talking about and, you know, feeling comfortable to talk to* [the therapist] *because I feel like a few more sessions would have been good. (*Ppt 4, Female). Expanding on this suggestion, we discussed the spacing of sessions and asked:

would offering a longer duration of therapy yield any additional benefits? However, the participants favoured the weekly spacing of the current design:

> *I think if you spread stuff out too much then you kind of lose the kind of train of thought. I guess there’s a more of a struggle to connect stuff from different sessions and if I thought something, like, on Monday and I had to wait all the way until, like, Saturday to tell* [the therapist], *that’s already a long time. But if it was, like, two weeks from then I just think it’s, like, too long and there’s not really a lot of opportunity to address immediate problems that you’re facing*. (Ppt2, Female)
>
> *I think the timing was about right*. (Ppt 7, Male)

During the clinical team consensus meeting, discussion concluded that a focus on increasing the amount, and the way that participants practice between sessions to integrate theory learnt during the therapy with life, could support a more intrinsic approach.

> *YP do need to be prepared to ‘act’* (e.g. accepting difficult feelings in the service of their values). *It’s not homework, its facilitating change for them* (tweak linguistically or the activities themselves to embed in their life not just about coming back to therapy or clinic appointments). (Psychologist)

However, feedback from the ‘Think-Aloud’ interviews suggested that additional work may not be favourably received:

> Interviewer: *Would you like a workbook to have alongside these sessions?*
>
> Ppt: *No, I actually don’t feel like I would use it*. (Ppt 12, Male, Session 7)

##### Outcomes

(i) The intervention was adapted to introduce discussions about eating, weight and self-image earlier, but with the scope to focus more intensely on it, throughout, being optional. For example, many activities request that the young person offers an example to work through, and the individual could choose to focus on an eating behaviour, body image concern or a more general example for the exercises. **(ii)** The programme structure of exercises that support rapport building within the early sessions was retained, and further rapport building exercises were made available for later sessions, if required to support the ongoing maintenance of a strong patient-therapist relationship. (iii) Online delivery modality retained, with space at the hospital offered for young people unable to have a private space at home to ensure inclusivity (this offer was available but not taken up during the current research). (iv) One-to-one therapy to be recommended as the first-choice approach, both directly answering the co-developers’ requests, and enabling the flexible manual approach. (v) To retain the young people’s ability to choose parental/companion involvement. (vi) The length of the therapy programme and the spacing between sessions would remain the same. However, there will be an increase to the expectation setting at the beginning of the programme to clarify how time with the therapist and independent time in the week to be implementing/practising the skills, come together to develop the outcome.

#### Qualitative theme: Views on strategies and engagement

The qualitative interviews facilitated feedback from the young people on a strategy-by-strategy basis. These data were used in real-time to evolve the therapy manual and were included in the Table of Changes (S1 Table).

An example of this feedback:

> *Our bad moments are like thunder or something, but it’s trying to remind ourselves that despite that thunder, that bad weather, that there’ll always be that good bit of sky there. It was one of the best ones* … *I didn’t really understand it at first, but after I thought about it a bit more it made more sense to me and I think, as I think about that more often, and I put that in practical situations, it’ll become more useful*. (Ppt 7, Session 5, Male)

One young person felt that the sessions were offering a different approach from the main clinic, but that sometimes this felt confusing or that the approaches conflicted with one another.

> *I mean, the fact that you’re at the CoCo clinic and kind of being told to kind of count calories and look at the diet and, you know, really focus on the food and take the drugs and so on, with the very clear focus about, you know, losing weight and focusing on that. And then in the counselling* [AIM2Change], *it’s much more about feeling good about yourself, being positive and it’s those two things are almost in conflict with each other*. (Ppt 2, Female)

It was suggested that a key outcome of the therapy could be seen to be that attending the therapy helps the young person to navigate the CEW clinics, and the difficult feelings and thoughts that arise during them.

> *How YP may feel when they are weighed* (using ACT to understand the world, clinic or family factors are part of that). (Psychologist)

##### Outcomes

(i) Data collected on individual strategies were used to adapt the manual. This process enabled the young people to shape the programme of therapy. (ii) Each therapy session should include a check-in regarding the young person’s experiences within the main CEW service that week, to ensure that they are being directly supported to understand any seemingly conflicting approaches and any difficult feelings that may have arisen.

#### Qualitative theme: Real world benefits of co-development process

The young people’s reflections suggest that they experienced benefits from taking part in this therapy. For instance, by incorporating strategies into their lives:

> *I think urge surfing or surfing the urge [strategy]. I happen to like ride along with it because there’s no point trying to stop a wave because it would just be wasting energy and have a more negative effect*. (Ppt 7, Session 6, Male)

Becoming more aware of their internal experiences:

> *As the sessions have gone on, it’s been easier to think about these sort of things. These analogies have been more clear. And with, like, the time going on, it’s much more easier to think about feelings or easy examples to actually give*. (Ppt 7, Session 3, Male)

And feeling that they were able to connect and share more with other people in their social world:

> *Well, I’m opening up to people more. I’m talking more about me and how much will work personal*ly *in my life. I thought, I really want to tell my friends I lost weight. Yeah*. (Ppt 8, Session 7, Male)

##### Outcome

The co-developers report anecdotal positive effects from experiencing the therapy that should be explored quantitatively in future research trials.

## Discussion

This paper reports the co-development of AIM2Change, in which young people attending a tier 3 weight management clinic received, and guided the development of, an ACT-based therapy. Overall, this co-development process, and the young people’s view therein, has provided preliminary evidence that an ACT/DNA-V approach is suitable for use in a paediatric weight management setting. In common with the wider paediatric weight management population, several of our co-developers experienced neurodiversity. Our findings are consistent with previous studies that indicate that ACT shows promise for individuals with ASD or ADHD(30, 31). The co-developers demonstrated a good comprehension of the model throughout, and even co-developers who were initially hesitant reported experiencing value in the model and therapeutic relationship during the study.

The multifaceted co-development process yielded valuable insights, with some clear changes that were unanimously agreed and simple to implement, such as the suggestion to enhance the opportunities for self-reflection or to tailor the more generic videos and activities to the individual’s context. Tailoring the structure and content of the sessions to individuals’ characteristics and preferences is known to be important to outcomes(32), and not meeting participants’ individual needs and expectations is a common reason for dropping out of weight management treatments in children(33).

The more complex decisions were discussed during the consensus meetings. Co-developers consistently recommended more therapy sessions, but as the intervention specifically sought to increase self-determination and reduce reliance of clinicians, it was pertinent to readdress this request. Consensus meeting concluded that instead of facilitating further clinician time, adaptions to the therapy were made to enhance self-determination. For example, ensuring from the beginning that patients were encouraged to think about themselves as having the power to make changes and that the therapist was here to guide the process. Greater emphasis was given during sessions and ‘homework’ tasks to apply the learning to examples in the young person’s own context, and the importance of using the skills outside of the therapy sessions was further embedded. The final session was adapted to have a ‘stronger exit strategy’, including a greater emphasis on reviewing the progress made by the young person, build self-confidence for independent work, and detailing practical ways in which they could continue to implement learnings in their life.

The consensus meetings also discussed an issue around the lack of consistency between the approach of AIM2Change and the focus of behaviour change offered by the CoCO clinic. The main CEW services are limited by time and limited psychological input. Lillis and Kendra(22) highlight potential barriers arising from the combination of ACT and standard behavioural therapy, referring to philosophical and structural contradictions such as non-aligned treatment goals, and ways of appraising thoughts and encouraging behavioural change. Nonetheless, literature suggests that the simultaneous application of both approaches may be feasible(22, 34). AIM2Change has the potential to give young people the time and space to reflect on frequently difficult conversations that they experience at the clinic, and to better understand how they experience their weight loss journey. It was also advised that the individual’s goals from the CoCo clinic be incorporated into the therapeutic process, to align outcomes, and support the young person to reach the goals set by clinicians in a sustainable, intrinsically motivated way.

### Strengths and limitations

Strengths of this research include the young co-developers’ positive attitude towards the therapist and the strong ‘therapeutic alliance’ that was built, with an online format. The alliance is considered essential as it provides an equal and cooperative footing in the process of change(32), which is aligned with both the ACT approach and with the intention to foster self-determination rather than a reliance on a therapeutic ‘expert’.

The flexible approach to both the content and timings of sessions was a strength that enabled participants to continue to participate throughout changeable life circumstances. Participants from a wide range of backgrounds, neurodiversity and readiness to engage in therapy were retained in this study. This is particularly noteworthy given that it has previously been indicated that young people from underserved communities (well represented in our co-developer sample) are more likely to disengage with paediatric weight management services(33).

The person-based approach, including co-development therapy sessions followed by consensus meetings with the young people, their parents, as well as clinicians and ACT experts, was deemed to be successfully applied by all contributors including the PPI advisory group. Indeed, this may be the first time the young people have engaged in such “meta-work” (i.e. experiencing and commenting on the therapeutic intervention at the same time). The young people were asked to distinguish between the therapy itself and their reflections and opinions on that therapy; this study has shown that this process is both possible and successful.

Whilst this study was centred on co-design of a new intervention, participants did appear to experience personal benefits from participating. Some co-developers suggested changes in the way they appraised situations, which increased their self-worth and acceptance, in line with the findings of previous studies(35, 36). Young people also reported improvements in school engagement and family and social interactions, consistent with Tronieri et al.(37), where benefits in family relations and social life-related quality of life were observed. The young people generalised their positive experiences, expecting others like them to feel the same way, which offers promise for the outcomes of a future randomised controlled trial. Members of the PPI advisory group found benefits in joining the project and agreed that contributing to the project had been a positive experience. They stated that they had been alerted to a new way of thinking about managing their weight, whilst feeling listened to and valued.

## Conclusion

The co-developers approved of the use of the ACT DNA-V model, tailored, jointly with them, for paediatric weight management in AIM2Change, within this population. Plans for future work include a pilot study to test the feasibility of the co-developed intervention across two CEW clinics, followed by an RCT in many CEW clinics in the UK to evaluate the effectiveness of the intervention. The invaluable insights provided by our co-developers and PPI members throughout this process have shaped and guided the intervention to ensure it best meets the specific needs of this population.

## Data Availability

All data produced in the present study are available upon reasonable request to the authors, and will be made available on data.bris on publication.

## Acknowledgments

We thank the young people and their parents who participated in the AIM2Change co-development project. We thank members of the steering group for their input into this study (Dr N. Davis, Mr R. Courtney-Tucker, Miss L. Stancheris). We also thank the members of the Patient and Public Advisory Group for their invaluable contributions throughout the project. We would also like to acknowledge the support of Professor A. Gibson and the CUBE team for facilitating the PPI evaluation, and Dr D. Giri and Dr K. Hawton during the recruitment stage of the study, and Dr E. Maiz as our valued collaborator.

## Supporting information

**S1 File. Inclusion questionnaire**.

**S1 Table. Table of changes**.

## References

1. (NCD-RisC) NRFC. Worldwide trends in body-mass index, underweight, overweight, and obesity from 1975 to 2016: a pooled analysis of 2416 population-based measurement studies in 128.9 million children, adolescents, and adults.. Lancet. 2017;390:2627–42.

2. World Health Organization. Obesity and overweight. 2024 [Available from: https://www.who.int/news-room/fact-sheets/detail/obesity-and-overweight.

3. Di Cesare M, Sorić M, Bovet P, Miranda J, Bhutta Z, Stevens GA, et al. The epidemiological burden of obesity in childhood: a worldwide epidemic requiring urgent action.. BMC Medicine 2019;17:212.

4. Pulgaron ER. Childhood obesity: a review of increased risk for physical and psychological comorbidities. Clin Ther. 2013;35(1):A18–32.

5. Hawton K, Apperley L, Parkinson J, Owens M, Semple C, Canvin L, Holt A, Easter S, Clark K, Lund K, Clarke E, O’Brien J, Giri D, Senniappan S, Shield JPH. Complications of excess weight seen in two tier 3 paediatric weight management services: an observational study. Arch Dis Child. 2025 Feb 19;110(3):216–220. doi: 10.1136/archdischild-2024-327286.

6. Darnton-Hill I, Nishida C, James WP. A life course approach to diet, nutrition and the prevention of chronic diseases. Public Health Nutr. 2004;7(1A):101–21.

7. Mandal S, Leary SD, Timpson N, Abeysekera K, Shield JPH. Associations of waist circumference to height ratio and body mass index through childhood and adolescence on blood pressure and risk of young adult hepatic steatosis: a cohort study. Arch Dis Child. 2025 May 26:archdischild-2024-328140. doi: 10.1136/archdischild-2024-328140.

8. Park MH, Sovio U, Viner RM, Hardy RJ, Kinra S. Overweight in childhood, adolescence and adulthood and cardiovascular risk in later life: pooled analysis of three british birth cohorts. PLoS One. 2013;8(7):e70684.

9. Public Health England. Health matters: obesity and the food environment. 2017 [Available from: https://www.gov.uk/government/publications/health-matters-obesity-and-the-food-environment/health-matters-obesity-and-the-food-environment--2.

10. National Institute for Health and Care Excellence. Obesity: identification, assessment and management. 2014 [Available from: https://www.nice.org.uk/guidance/cg189.

11. Gungor NK. Overweight and obesity in children and adolescents. J Clin Res Pediatr Endocrinol. 2014;6(3):129–43.

12. https://www.gov.uk/government/publications/childhood-obesity-a-plan-for-action/childhood-obesity-a-plan-foraction#:~:text=As%20a%20first%20major%20step,in%20the%20Finance%20Bill%202017.13.

13. https://efaidnbmnnnibpcajpcglclefindmkaj/ https://www.instituteforgovernment.org.uk/sites/default/files/2023-04/tackling-obesity.pdf

14. Apperley LJ, Blackburn J, Erlandson-Parry K, Gait L, Laing P, Senniappan S. Childhood obesity: A review of current and future management options. Clin Endocrinol (Oxf). 2022;96(3):288–301.

15. Cox JS, Searle AJ, Hinton EC, Giri D, Shield JPH. Perceptions of non-successful families attending a weight-management clinic. Arch Dis Child. 2021;106(4):377–82.

16. Gillison FB, Standage M, Skevington SM. Relationships among adolescents’ weight perceptions, exercise goals, exercise motivation, quality of life and leisure-time exercise behaviour: a self-determination theory approach. Health Educ Res. 2006;21(6):836–47.

17. Jensen CD, Duraccio KM, Hunsaker SL, Rancourt D, Kuhl ES, Jelalian E, et al. A qualitative study of successful adolescent and young adult weight losers: implications for weight control intervention. Child Obes. 2014;10(6):482–90.

18. Cox JS, Searle A, Thornton G, Hamilton-Shield JP, Hinton EC. Integrating COM-B and the person-based approach to develop an ACT based therapy programme to raise selfdetermination in adolescents with obesity. BMC Health Serv Res. 2023;23(1):1158.

19. Hayes SC, Luoma JB, Bond FW, Masuda A, Lillis J. Acceptance and commitment therapy: model, processes and outcomes. Behav Res Ther. 2006;44(1):1–25.

20. Lawlor ER, Islam N, Bates S, Griffin SJ, Hill AJ, Hughes CA, et al. Third-wave cognitive behaviour therapies for weight management: A systematic review and network metaanalysis. Obes Rev. 2020;21(7):e13013.

21. Iturbe I, Echeburua E, Maiz E. The effectiveness of acceptance and commitment therapy upon weight management and psychological well-being of adults with overweight or obesity: A systematic review. Clin Psychol Psychother. 2022;29(3):837–56.

22. Lillis J, Kendra KE. Acceptance and commitment therapy for weight control: Model, evidence, and future directions. J Contextual Behav Sci. 2014;3(1):1–7.

23. Cox JS, Iturbe I, Searle A, Maiz E, Hinton EC. The acceptability, feasibility and preliminary efficacy of acceptance and commitment therapy for adolescents in the management of overweight or obesity: A scoping review. Journal of Contextual Behavioral Science. 2024;31(100713).

24. Hayes LL, Ciarrochi JV. The thriving adolescent: Using acceptance and commitment therapy and positive psychology to help teens manage emotions, achieve goals, and build connection. : New Harbinger Publications; 2015.

25. Yardley L, Morrison L, Bradbury K, Muller I. The person-based approach to intervention development: application to digital health-related behavior change interventions. J Med Internet Res. 2015;17(1):e30.

26. https://www.nihr.ac.uk/research-funding/application-support/working-with-people-and-communities

27. Gibson A, Welsman J, Britten N. Evaluating patient and public involvement in health research: from theoretical model to practical workshop. Health Expectations. 2017;20:826–35.

28. Hinton EC, Fenwick C, Hall M, Bell M, Hamilton-Shield JP, Gibson A. Evaluating the benefit of early patient and public involvement for product development and testing with small companies. Health Expect. 2023;26(3):1159–69.

29. Gale NK, Heath G, Cameron E, Rashid S, Redwood S. Using the framework method for the analysis of qualitative data in multi-disciplinary health research. BMC Med Res Methodol. 2013;13:117.

30. Pahnke J, Hirvikoski T, Bjureberg J, Bölte S, Jokinen J, Bohman B, et al. Acceptance and commitment therapy for autistic adults: An open pilot study in a psychiatric outpatient context. Journal of Contextual Behavioral Science. 2019;13:34–41.

31. Vanzin L, Mauri V, Valli A, Pozzi M, Presti G, Oppo A, et al. Clinical Effects of an ACT-Group Training in Children and Adolescents with Attention-Deficit/Hyperactivity Disorder. Journal of Child and Family Studies. 2019;29(4):1070–80.

32. Walser RD, O’Connell M. Acceptance and commitment therapy and the therapeutic relationship: Rupture and repair. J Clin Psychol. 2021;77(2):429–40.

33. Skelton JA, Beech BM. Attrition in paediatric weight management: a review of the literature and new directions. Obes Rev. 2011;12(5):e273–81.

34. Forman EM, Butryn ML. A new look at the science of weight control: how acceptance and commitment strategies can address the challenge of self-regulation. Appetite. 2015;84:171–80.

35. Cardel MI, Lee AM, Chi X, Newsome F, Miller DR, Bernier A, et al. Feasibility/acceptability of an acceptance-based therapy intervention for diverse adolescent girls with overweight/obesity. Obes Sci Pract. 2021;7(3):291–301.

36. Newsome FA, Cardel MI, Chi X, Lee AM, Miller D, Menon S, et al. Wellness Achieved Through Changing Habits: A Randomized Controlled Trial of an Acceptance-Based Intervention for Adolescent Girls With Overweight or Obesity. Child Obes. 2023;19(8):525–34.

37. Tronieri JS, Wadden TA, Leonard SM, Berkowitz RI. A pilot study of acceptance-based behavioural weight loss for adolescents with obesity. Behav Cogn Psychother. 2019;47(6):686–96.

